# National Routine Adult Immunization Programs among World Health Organization Member States: An Assessment of Health Systems to Deploy Future SARS-CoV-2 Vaccines

**DOI:** 10.1101/2020.10.16.20213967

**Authors:** Sarah R. Williams, Amanda J. Driscoll, Hanna M. LeBuhn, Wilbur H. Chen, Kathleen M. Neuzil, Justin R. Ortiz

## Abstract

**Introduction:** As the SARS-CoV-2 pandemic disproportionately affects older adults, future pandemic vaccine response will rely on existing adult immunization infrastructures.

**Methods:** We evaluated the 2018 WHO/UNICEF Joint Reporting Form on Immunization for country reports on adult immunization programs. We described countries with programs and used multivariable regression to identify independent factors associated with having them.

**Results:** Of 194 WHO Member States, 120 (62%) reported having any adult vaccination program. The Americas and Europe had the most adult immunization programs, most commonly Hepatitis B and influenza vaccines (>45% and >90% of countries). Africa and South-East Asia had the fewest adult immunization programs, with <11% of countries reporting any adult immunization programs for influenza or hepatitis vaccines, and none for pneumococcal vaccines. In bivariate analyses, high- or upper-middle income, introduction of new or underused vaccines, having achieved pediatric vaccine coverage goals, and meeting National Immunization Technical Advisory Groups basic functional indicators were significantly associated (p<0.001) with having any adult immunization programs. In multivariable analyses, the factor most strongly associated with adult immunization programs was country income, with high- or upper-middle income countries significantly more likely to report having a program (aOR 19.3, 95% CI 6.5, 57.7).

**Discussion:** That 38% of countries lack functional platforms for adult immunization has major implications for future SARS-CoV-2 vaccine deployment. Systems for vaccine storage and handling, delivery, and waste management for adult immunization do not exist in much of the world. Developing countries should strengthen immunization programs to reach adults with SARS-CoV-2 vaccines when they become available.

## INTRODUCTION

The global community is rapidly mobilizing to develop and license vaccines to prevent SARS-CoV-2 infection. While the timely development and manufacture of SARS-CoV-2 vaccines is a public health imperative, vaccines will have to be deployed rapidly and globally to provide the greatest impact. Health care system workers are at high-risk of exposure to SARS-CoV-2 [1], and maintaining essential health services is critical during a pandemic. Likewise, older adults and adults with chronic medical conditions are at the greatest risk of severe SARS-CoV-2 disease [1]. Any global pandemic vaccine response will rely on existing immunization infrastructures to reach adult target groups [2].

While the World Health Organization (WHO) recommends several vaccines for use by adults, global immunization efforts have traditionally focused on young children. Since 1974, the Expanded Programme on Immunization (EPI) has been the major platform for vaccine delivery in low-income countries (LICs) and lower-middle-income countries (LMICs) [3]. Built on the global smallpox eradication infrastructure, EPI originally included four vaccines against six infectious diseases administered in the first year of life: bacille Calmette-Guerin, diphtheria-tetanus-pertussis, polio, and measles vaccines [3]. Since then, EPI has been expanded to include several additional vaccines, with most vaccines still targeting children in the first two years of life [4]. Exceptions include maternal tetanus toxoid vaccination in settings where routine paediatric tetanus immunization is suboptimal and where maternal and neonatal tetanus have not been eliminated [5] and human papillomavirus vaccines (HPV) targeting girls aged nine to 14 years [6]. WHO also recommends additional vaccines for adults in regions where certain diseases are endemic (such as Japanese encephalitis and yellow fever vaccines), in high-risk populations (such as influenza and rabies vaccines), and under special circumstances (such as in outbreak settings with vaccines against Ebola virus disease, cholera, or yellow fever) [4].

To better understand the status of national immunization programs targeting adults globally, we reviewed 2018 adult immunization data reported by countries through the Joint Reporting Form on Immunization (JRF) to WHO and UNICEF. We chose five routine vaccines licensed for adult immunization: hepatitis B vaccine (HepB), herpes zoster vaccine (HZV), influenza vaccine, pneumococcal conjugate vaccine (PCV), and pneumococcal polysaccharide vaccine (PPSV) (Table 1). We chose not to review adult programs for booster doses of routine childhood vaccines, vaccines that target regional endemic infections, or those that are used primarily in outbreak settings. Our objectives were to identify countries with routine adult immunization programs, to determine factors associated with having those programs, and to identify factors that could be addressed in order to strengthen the SARS-CoV-2 pandemic vaccine response.

**Table 1.** Summary of WHO Position Papers for Vaccine Use in Adults as of November 2020.

| Vaccine | Position Statement Year | Position for Vaccine Use in Adults |
| --- | --- | --- |
| Hepatitis B vaccine [26] | 2017 | The Position Paper states: “WHO recommends hepatitis B vaccination of persons at high-risk of hepatitis B virus infection in [adults] and catch-up vaccination of unvaccinated cohorts if the necessary resources are available.” |
| Herpes zoster vaccine [27] | 2014 | Countries that have an aging population and elevated disease burden may choose to introduce HZV. Citing unknown burden of disease and insufficient data supporting HZV in most countries, the WHO does not offer any recommendation regarding the routine use of HZV. |
| Influenza vaccine [28] | 2012 | The Position Paper states: “[I]ndividual national decisions on the use of influenza vaccines will be determined by national capacity and resources... For countries considering the initiation or expansion of programs for seasonal influenza vaccination, WHO recommends that pregnant women should have the highest priority. Additional risk groups to be considered for vaccination, in no particular order of priority, are children aged 6–59 months, [older adults], individuals with specific chronic medical conditions, and health-care workers.” |
| Pneumococcal conjugate vaccine [4, 29] | N/A | WHO does not currently have recommendations on the use of PCV in individuals over 5 years of age, although addressing pneumococcal vaccination in adults is on the WHO Strategic Advisory Group of Experts on Immunization (SAGE) agenda for future deliberation. |
| Pneumococcal polysaccharide vaccine [30] | 2012 | The Position Paper states: “Many industrialized countries recommend PPSV immunization of older adults and other high-risk groups. In resource-limited settings where there are many competing health priorities, the evidence does not support routine immunization of older adults and high-risk populations with PPSV. Given the substantial effects of herd immunity in adult age groups following routine infant immunization with PCV, a higher priority should be given to introducing and maintaining high coverage of infants with PCV. Countries considering introducing PPSV to [older adults] or other high-risk populations will need to develop strategies for reaching these target populations...The optimal timing, frequency and clinical effectiveness of additional doses of PPSV are poorly defined, and national recommendations regarding revaccination vary. However, on the basis of the data on the duration of vaccine-induced protection, WHO suggests one single revaccination >5 years after a first vaccination.” |

Abbreviations: herpes zoster vaccine=HZV, pneumococcal conjugate vaccine=PCV, pneumococcal polysaccharide vaccine=PPSV.

## METHODS

### Primary data source

We collected data on national adult immunization programs from the JRF [7]. The JRF is a monitoring and evaluation tool that collects national administrative information regarding estimates of immunization coverage, reported cases of vaccine-preventable diseases, immunization schedules, and vaccination campaigns, as well as indicators of immunization system performance and financing [8]. The JRF is the only database of its type to include country-level data on immunization programs globally, making it the only dataset to our knowledge that would permit a global analysis of adult immunization programs. We accessed the 2018 JRF database on 12 February 2020, and collected country data on the presence of adult immunization programs for HepB, HZV, influenza vaccine, PCV, and PPSV. To assess whether countries are adopting global mandates for new vaccine introductions, we identified countries that had introduced rotavirus vaccine, HPV, or HepB. Rotavirus vaccine and HPV vaccines are considered “new or underutilized” and their introduction is prioritized by the Global Vaccine Action Plan (GVAP). Birth dose HepB is recommended by WHO, but has lower coverage than most other vaccines in the EPI schedule, so we also included birth dose HepB as an indicator for adoption of recommended vaccination programs. To assess the general strength of routine immunization, we classified countries according to whether they had achieved the GVAP goals of maternal and neonatal tetanus elimination (less than one case of neonatal tetanus per 1,000 live births in every district of a country) [9] and whether they had achieved ≥95% national coverage of the third dose of diphtheria-tetanus-pertussis containing vaccine (DTP3) per WHO/UNICEF estimates [10]. To evaluate the capacity to make national decisions about vaccination programs, we used WHO data indicating whether countries had achieved WHO-defined process indicators for National Immunization Technical Advisory Groups (NITAGs) [9, 11]. NITAGs achieving these indicators (called “functional NITAGs”) have the following characteristics: 1) legislative or administrative basis for the advisory group; 2) formal written terms of reference; 3) at least five different areas of expertise represented among core members; 4) at least one meeting per year; 5) circulation of the agenda and background documents at least one week prior to meetings, and; 6) mandatory disclosure of any conflict of interest.

Questions in the 2018 JRF asked whether a country had adult programs for influenza vaccine, HepB, HZV, or PPSV. No specific questions were included for adult PCV programs, but there were questions about which groups were targeted for immunization with PCV with free-text fields for responses. For PCV, when it was clear that adult age groups were targeted by a particular vaccination program (such as when “adults,” “health care workers,” or persons aged “>65 years” were indicated), we considered that the country had reported affirmatively that an adult program was present. For countries that did not indicate the presence or absence of an adult immunization program in the JRF or that did not make clear whether an adult group was targeted for vaccination (such as when “high-risk groups” was indicated), we considered that the country did not report that an adult program was present.

### Other data sources

We collected additional information to supplement the JRF data. To the extent possible, all covariate data were also from 2018, the year of the JRF dataset. We sought relevant country immunization program information from websites of WHO Regional Offices, and we requested information from immunization staff in each WHO Regional Office. Regional Office immunization focal points are well-informed about routine immunization programs in their regions, they are a part of the annual JRF data quality review, and we considered their concurrence to country reports to be an important quality check for this project. We also accessed immunization schedules for countries in the European Union from the European Centers for Disease Control (ECDC) website [12]. Next, we collected country economic information. As low-resource countries often rely on financial support from Gavi (the Vaccine Alliance) for new vaccine introduction, we identified all countries eligible for Gavi funding support in 2018 [13]. For per capita health expenditures, we used World Bank data for 2016, and we classified country income categories using the 2018 World Bank designations [14, 15]. For population analyses, we used 2017 population estimates from the Institute for Health Metrics and Evaluation [16].

### Statistical analysis

Because countries are not required to record the absence of immunization programs in the JRF for all the vaccines we analysed, we could only record affirmative responses for some vaccines. For analyses, we used the number of relevant WHO Member States as denominators because we were unable to distinguish country non-responses from negative responses. Further, PCV program questions are not specific for adults. As many countries indicate presence of paediatric pneumococcal vaccination programs, we could not simply exclude countries from a denominator if they did not respond to a pneumococcal vaccination program question. For statistical analyses, income group was defined as a categorical variable, the presence of adult immunization programs and health system/immunization program indicators were defined as dichotomous variables, and per capita health expenditure was defined as a continuous variable. We used descriptive statistics to describe countries with adult immunization programs. We conducted bivariate analyses to determine whether certain health system/immunization program characteristics were associated with the reported presence of any adult immunization program using χ2 tests for categorical variables and Kruskal-Wallis tests for comparisons of medians. We calculated p-values for trends using an extension of the Wilcoxon rank-sum test and adjusted odds ratios (aOR) using multiple logistic regression. The bivariate and multiple logistic regression analyses were performed on the complete global dataset and specifically for the European Region. All statistical tests were two-sided, and p-values <0.05 were considered statistically significant. Analyses were conducted in Stata (version 15.1, StataCorp, College Station, Texas). Human participants were not involved in this study, so institutional review board approval was not required.

## RESULTS

### Data missingness

Among the 194 WHO Member States, 176 (90.7%) reported the presence or absence of a national adult immunization program in the JRF for at least one of the five vaccines -- range by region: 6/11 (54.5%) in South-East Asia to 35/35 (100.0%) in the Americas. Responses were available for 83 (42.8%) countries on the presence of an adult vaccination program for HepB, 38 (19.6%) countries for HZV, 114 (58.8%) countries for influenza vaccine, 157 (80.9%) countries for PCV, and 59 (30.4%) countries for PPSV. Otherwise country JRF responses were missing (Supplemental Table 1). Our review of ECDC country policy information identified 20 vaccine policies that were not reported in the 2018 JRF. Supporting information subsequently obtained from WHO Regional websites and Regional Office immunization program officers did not identify any new or conflicting data. Percentages of countries with particular policies are presented as the number of countries reporting or identified through additional efforts to have a particular vaccination programs as numerators, with all WHO Member States by geographic category as denominators.

### Programs globally and by region

Among the 194 WHO Member States, 71 (36.7%) countries reported having adult immunization programs for HepB, 15 (8.8%) countries for HZV, 114 (58.8%) for influenza vaccine, 16 (8.3%) for PCV, and 35 (18.0%)for PPSV (Table 2). A total of 120 (61.9%) countries reported having any of the five assessed adult immunization programs, while 3 (2.5%) countries reported having programs for each of the five vaccines studied (Greece, Italy, and the United States).

**Table 2.**
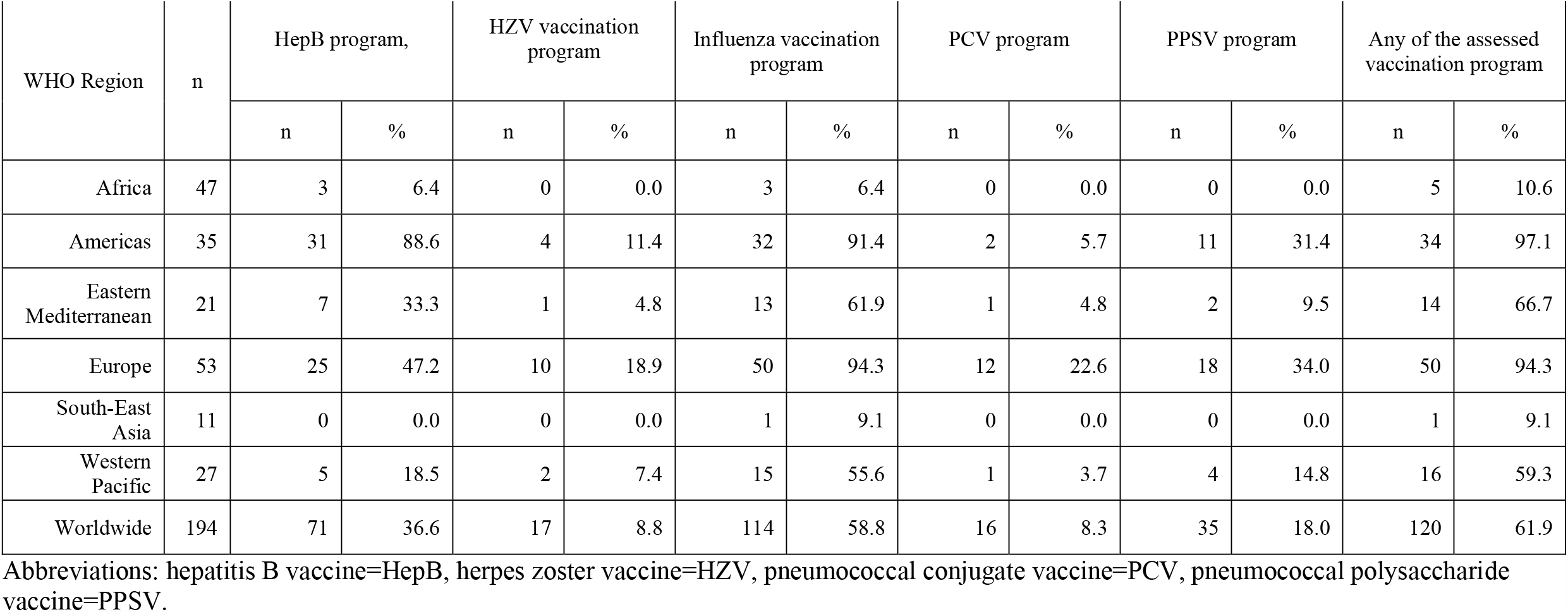
WHO Member States Affirmatively Reporting National Adult Immunization Programs in 2018, by WHO Region and Globally.

| WHO Region | n | HepB program, |  | HZV vaccination program |  | Influenza vaccination program |  | PCV program |  | PPSV program |  | Any of the assessed vaccination program |  |
| --- | --- | --- | --- | --- | --- | --- | --- | --- | --- | --- | --- | --- | --- |
|  |  | n | % | n | % | n | % | n | % | n | % | n | % |
| Africa | 47 | 3 | 6.4 | 0 | 0.0 | 3 | 6.4 | 0 | 0.0 | 0 | 0.0 | 5 | 10.6 |
| Americas | 35 | 31 | 88.6 | 4 | 11.4 | 32 | 91.4 | 2 | 5.7 | 11 | 31.4 | 34 | 97.1 |
| Eastern Mediterranean | 21 | 7 | 33.3 | 1 | 4.8 | 13 | 61.9 | 1 | 4.8 | 2 | 9.5 | 14 | 66.7 |
| Europe | 53 | 25 | 47.2 | 10 | 18.9 | 50 | 94.3 | 12 | 22.6 | 18 | 34.0 | 50 | 94.3 |
| South-East Asia | 11 | 0 | 0.0 | 0 | 0.0 | 1 | 9.1 | 0 | 0.0 | 0 | 0.0 | 1 | 9.1 |
| Western Pacific | 27 | 5 | 18.5 | 2 | 7.4 | 15 | 55.6 | 1 | 3.7 | 4 | 14.8 | 16 | 59.3 |
| Worldwide | 194 | 71 | 36.6 | 17 | 8.8 | 114 | 58.8 | 16 | 8.3 | 35 | 18.0 | 120 | 61.9 |

Abbreviations: hepatitis B vaccine=HepB, herpes zoster vaccine=HZV, pneumococcal conjugate vaccine=PCV, pneumococcal polysaccharide vaccine=PPSV.

Adult vaccination program data by WHO Region are in Figure 1 and Table 2. In the African Region (47 countries), 5 (10.6%) countries reported at least one adult vaccination program. Three (6.4%) countries reported HepB programs, and 3 (6.4%) countries reported influenza vaccination programs. The three countries reporting influenza vaccination programs in the African Region (Algeria, South Africa, and Mauritius) were not the same three reporting HepB programs (Eswatini, Kenya, and Mauritius). No African countries reported adult vaccination programs for HZV, PCV, or PPSV. In the Americas (35 countries), 34 (97.1%) countries reported at least one adult vaccination program. American countries had programs for PCV (2 countries; 5.7%), HZV (4 countries, 11.4%), PPSV (11 countries, 31.4%), HepB (31 countries, 88.6%), and influenza vaccine (32 countries, 91.4%). In the Eastern Mediterranean (21 countries), 14 (66.7%) countries reported at least one adult vaccination program. Eastern Mediterranean countries had programs for HZV and PCV (1 country each, 4.8%), PPSV (2 countries, 9.5%), HepB (7 countries, 33.3%), and influenza vaccine (13 countries, 61.9%). In the European Region (53 countries), 50 (94.3%) countries reported at least one adult vaccination program. European countries had programs for HZV (10 countries, 18.9%), PCV (12 countries, 22.6%), PPSV (18 countries, 34.0%), HepB (25 countries, 47.2%), and influenza vaccine (50 countries, 94.3%). In South-east Asia (11 countries), 1 (9.1%) country reported any adult vaccination program – an adult influenza vaccination program in Thailand. No other South-east Asian countries reported programs for HepB, HZV, PCV, or PPSV. In the Western Pacific (27 countries), 16 (59.3%) countries reported any adult vaccination program. Western Pacific countries had programs for PCV (1 country, 3.7%), HZV (2 countries, 7.4%), PPSV (4 countries, 14.8%), HepB (5 countries, 18.5%), and influenza vaccine (15 countries, 55.6%).

**Figure 1.**
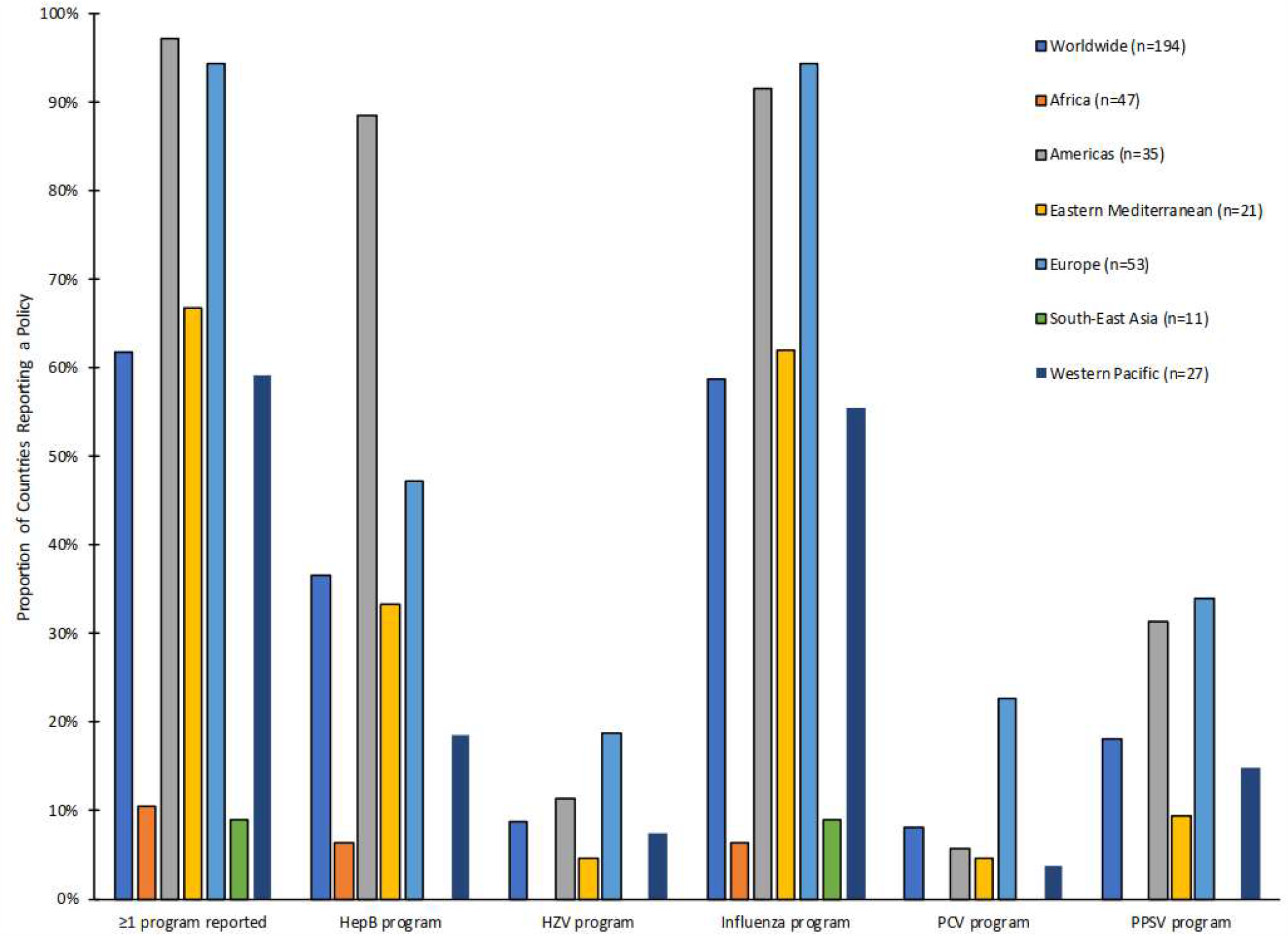
Reported Adult Immunization Programs by WHO Region in 2018. Note: From 2018 WHO/UNICER Joint Reporting Form [7]. Abbreviations: HepB = Hepatitis B vaccine; HZV= Herpes Zoster Vaccine; PCV = pneumococcal conjugate vaccine; PPSV = pneumococcal polysaccharide vaccine; HPV= human papillomavirus vaccine

We compared the populations of countries with and without adult vaccination programs. Globally, 38.7% of the world’s population lives in a country reporting any adult vaccination program (Table 3). The percentage of the world population living in countries reporting specific adult vaccination programs includes the following: 13.3% for HepB, 9.0% for HZV, 36.7% for influenza vaccine, 30.9% for PCV, and 19.1% for PPSV. The percentage of persons living in countries reporting any adult vaccination program by WHO Region is 13.9% for the African Region, 98.8% for the Americas, 51.2% for the Eastern Mediterranean Region, 97.2% for the European Region, 3.5% for the South-east Asian Region, and 24.1% for the Western Pacific Region.

**Table 3.** Population in Countries Reporting National Adult Immunization Programs in 2017, by WHO Region and Globally.

| WHO Region | Population<br>(in millions) | Population in countries that<br>report HepB program |  | Population in countries that<br>report HZV program |  | Population in countries that<br>report Influenza vaccination<br>program |  | Population in countries that<br>report PCV program |  | Population in countries that<br>report PPSV program |  | Population in countries that<br>report having any of the<br>assessed vaccination<br>programs |  |
| --- | --- | --- | --- | --- | --- | --- | --- | --- | --- | --- | --- | --- | --- |
|  |  | n (in millions) | % | n (in millions) | % | n (in millions) | % | n (in millions) | % | n (in millions) | % | n (in millions) | % |
| Africa | 1049.6 | 50.7 | 4.8 | 0 | 0.0 | 96.7 | 9.2 | 0 | 0.0 | 0 | 0.0 | 146.1 | 13.9 |
| Americas | 1002.9 | 686.2 | 68.4 | 412.0 | 41.1 | 990.2 | 98.7 | 328.3 | 32.7 | 640.4 | 63.9 | 991.0 | 98.8 |
| Eastern<br>Mediterranean | 706.1 | 157.9 | 22.4 | 9.7 | 1.4 | 265.3 | 37.6 | 93.4 | 13.2 | 18.2 | 2.6 | 361.8 | 51.2 |
| Europe | 932.5 | 79.6 | 8.5 | 231.7 | 24.8 | 906.6 | 97.2 | 160.5 | 17.2 | 476.7 | 51.1 | 906.6 | 97.2 |
| South-East Asia | 1999.0 | 0 | 0.0 | 0 | 0.0 | 70.6 | 3.5 | 0 | 0.0 | 0 | 0.0 | 70.6 | 3.5 |
| Western Pacific | 1896.1 | 36.0 | 1.9 | 28.4 | 1.5 | 456.0 | 24.0 | 4.4 | 0.2 | 314.0 | 16.6 | 456.9 | 24.1 |
| Worldwide | 7586.3 | 1010.4 | 13.3 | 681.8 | 9.0 | 2785.4 | 36.7 | 586.6 | 30.9 | 1449.4 | 19.1 | 2933.1 | 38.7 |

Abbreviations: hepatitis B vaccine=HepB, herpes zoster vaccine=HZV, pneumococcal conjugate vaccine=PCV, pneumococcal polysaccharide vaccine=PPSV.

In bivariate analyses, economic indicators were highly associated with having an adult immunization program (Figure 2 and Table 4). Countries classified by the World Bank as high- or upper-middle income were more likely to have any adult immunization program than those classified as low- or lower-middle income (p<0.001), and countries with any adult immunization program had higher median per capita health expenditures compared to those without a program (p<0.001), with the same being true for the individual adult vaccination programs (Supplemental Tables 2 and 3). Certain characteristics of the national immunization system were also associated with a reported adult immunization program in the JRF (Table 4). Compared to countries without any reported adult vaccination program, those countries that reported any adult vaccination programs were more likely to have included the HepB birth dose in their routine immunization schedule (94/120; 78.3%) vs. (34/74; 45.9%, p<0.001), to have eliminated maternal and neonatal tetanus (120/120; 100% vs. 60/74; 81.1%, p<0.001), and to have achieved national DTP3 coverage ≥95% (66/120; 55.0% vs. 18/74; 24.3%, p<0.001). Having introduced HPV was associated with having at least one adult vaccination program (76/120; 46.3% vs. 14/74; 18.9%, p<0.001) as well as with having each of the five individual adult vaccination programs (Supplemental Table 4). The introduction of rotavirus vaccine was not associated with the presence of an adult vaccination program (Table 3). Finally, countries reporting an adult vaccination program were more likely to have a functional NITAG than those countries not reporting an adult vaccination program (82/120; 68.3% vs. 32/74; 43.2%, p<0.001).

**Table 4.** Characteristics of WHO Member States in 2018 with and without any Reported Adult Immunization Program.

|  | Any reported adult vaccination program |  |  |  |  |
| --- | --- | --- | --- | --- | --- |
|  | Yes<br>N=120 |  | No<br>N=74 |  |  |
|  | Median (IQR) |  | Median (IQR) |  | p-value<sup>2</sup> |
| Median per capita health expenditure (USD)<sup>3</sup> | 854 (396, 2215) |  | 58 (22, 153) |  | <0.001 |
|  | n | % | n | % | p-value<sup>4</sup> |
| World Bank income group<sup>5</sup> |  |  |  |  |  |
| Low income | 1 | 0.8 | 33 | 45.2 | <0.001 |
| Lower-middle income | 18 | 15.1 | 28 | 38.4 |  |
| Upper middle income | 45 | 37.8 | 11 | 15.1 |  |
| High income | 55 | 46.2 | 1 | 1.4 |  |
| Gavi eligible |  |  |  |  |  |
| Yes | 1 | 0.8 | 47 | 63.5 | <0.001 |
| No | 119 | 99.2 | 27 | 36.5 |  |
| Introduced Hepatitis B vaccine birth dose |  |  |  |  |  |
| Yes | 94 | 78.3 | 34 | 45.9 | <0.001 |
| No | 26 | 21.7 | 40 | 54.1 |  |
| Introduced human papilloma virus vaccine |  |  |  |  |  |
| Yes | 76 | 63.3 | 14 | 18.9 | <0.001 |
| No | 44 | 36.7 | 60 | 81.1 |  |
| Introduced rotavirus vaccine |  |  |  |  |  |
| Yes | 60 | 50.0 | 41 | 55.4 | 0.46 |
| No | 60 | 50.0 | 33 | 44.6 |  |
| Functional NITAG |  |  |  |  |  |
| Yes | 82 | 68.3 | 32 | 43.2 | <0.01 |
| No | 38 | 31.7 | 42 | 56.8 |  |
| Eliminated maternal and neonatal tetanus |  |  |  |  |  |
| Yes | 120 | 100.0 | 60 | 81.1 | <0.001 |
| No | 0 | 0.0 | 14 | 18.9 |  |
| Third dose diphtheria-tetanus-pertussis coverage ≥95% nationally | | | | | |
| Yes | 66 | 55.0 | 18 | 24.3 | <0.001 |
| No | 54 | 45.0 | 56 | 75.7 |  |

Notes:
1. Adult immunization program for any of the following: hepatitis B vaccine, herpes zoster vaccine, pneumococcal conjugate vaccine, pneumococcal polysaccharide vaccine, or human papilloma virus vaccine 2. Kruskal-Wallis test for difference in medians 3. Excluding 11 countries with missing health care expenditure data 4. P-value for trend (World Bank income group comparisons) and chi-square test (other comparisons) 5. Denominator =192 countries; 119 with an adult vaccine program and 73 without an adult vaccine program. Niue and The Cook Islands, not World Bank member countries, are excluded from the income categories. All data are from 2018 except per capita health expenditures, which are from 2016, the most recent year for which such data were available from the World Bank.

**Figure 2.**
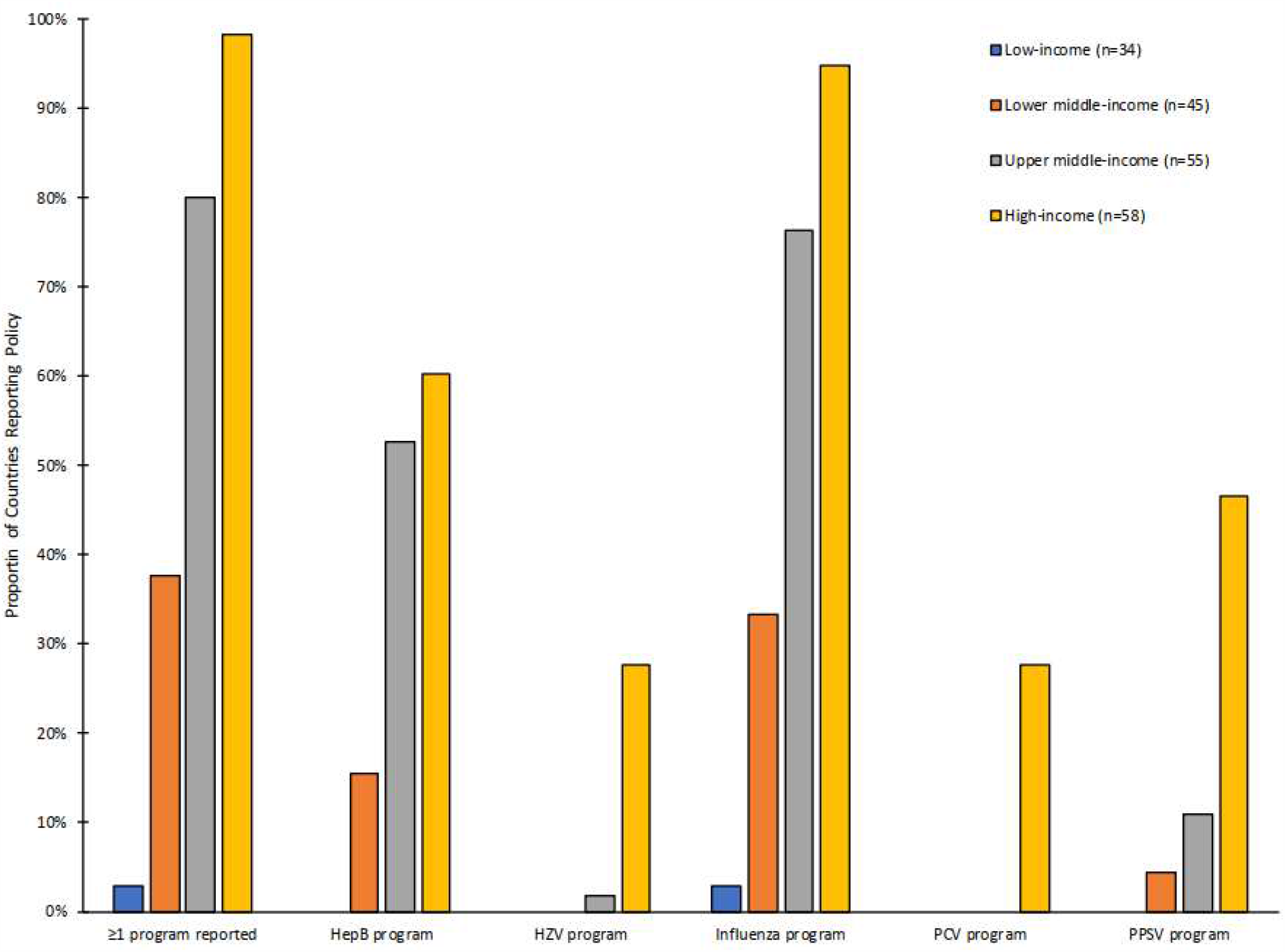
Reported Adult Immunization Programs by World Bank Income Category in 2018. Note: From 2018 WHO/UNICEF Joint Reporting Form [31]. Abbreviations: HepB = Hepatitis B vaccine; HZV= Herpes Zoster Vaccine; PCV = pneumococcal conjugate vaccine; PPSV = pneumococcal polysaccharide vaccine; HPV= human papilloma virus vaccine

In a multiple logistic regression model including each of the binary national immunization system indicators (≥95% DTP3 coverage, presence of a functioning NITAG, HepB birth dose, rotavirus vaccine, and HPV vaccine), and adjusting for income category, (LIC/LMIC vs. upper-middle-income country [UMIC]/ high-income country [HIC]), the presence of a functional NITAG (aOR 7.1, 95% confidence interval (CI) 2.5, 20.7), the use of the HepB birth dose (aOR 3.6, 95% CI 1.4, 10.4), and the use of HPV (aOR 3.9, 95% CI 4.3, 10.4) remained significantly associated with reporting an adult immunization program (Supplemental Table 5). Country income status was the factor most strongly associated with reporting an adult immunization program (aOR 21.9, 95% CI 7.3, 65.3) for HIC/UMIC countries compared to LIC/LMIC countries. In a model limited to WHO Member States classified as LIC or LMIC, the aOR for any adult immunization program was 12.0 (95% CI 2.3, 63.4) for countries with a functional NITAG compared to those without, and the aOR for any adult immunization program was 5.9 (95% CI 1.5, 24.2) for countries that had included birth dose HepB in the EPI schedule compared to those that had not. Multiple logistic regression models for the individual adult vaccination programs showed similar trends (Supplemental Table 6).

To gain insight into adult immunization programs in the WHO European Region, we conducted bivariate and multiple logistic regression analyses restricted to the 53 countries in that region. Of the 53 countries, three had no adult vaccination program (Azerbaijan, Kyrgyzstan and Tajikistan). In bivariate analyses, countries with at least one adult vaccination program had greater median per capita health expenditures compared to those without any adult program ($1,616 vs. $93, p<0.01 and were less likely to be Gavi eligible (0/50; 0% vs. 2/3; 66.7%, <0.001) (Supplemental Table 7). Having introduced an HPV vaccine was the only immunization program characteristic associated with having one or more adult vaccination programs (35/50; 70.0% vs. 0/3; 33.3%, p=0.01). In multivariable analyses adjusting for rotavirus introduction, having a functional NITAG, having DTP coverage ≥95% and HIC or UMIC country status), the only characteristic significantly associated with having an adult vaccination program was being a HIC or UMIC country (45/50; 90% vs. 1/3; 33.3%, aOR=37.3, 95% CI 1.5, 962.2) (Supplemental table 7). In multivariable analyses of the individual adult vaccination programs, having introduced HPV was independently associated with having an adult HepB program (20/25; 80% vs. 15/28; 53.6%, aOR=5.6, 95% CI 1.3, 24.2) and with having an adult PPSV program (17/18 94.4% vs. 18/35; 51.4%, aOR=11.7, 1.2, 112.4). In the European Region, being a HIC or UMIC, rather than a LMIC, was independently associated with having an adult influenza vaccination program (45/50; 90% vs. 1/3 33.3%, aOR=37.3 (1.4 vs. 962.2) (Supplemental Table 8).

## CONCLUSIONS

In our review of the 2018 JRF, 61.9% of WHO Member States reported having any adult vaccination programs, including countries from all WHO Regions and income categories. The most common adult immunization program was for influenza vaccination, reported by 58.8% of countries, while adult immunization programs for other vaccines were much less common. The number of adult influenza immunization programs has increased from an analysis of the 2014 JRF, when 46% of countries reported programs for adults with chronic disease and 45% reported programs for older adults [8]. In that analysis, countries reporting influenza immunization programs were wealthier, and they were more likely to have functional NITAGs, to have introduced new or under-utilized vaccines, and to have stronger immunization systems. Our study documents and quantifies major limitations to routine adult immunization programs globally and indicates that the trends seen for influenza vaccination are also true for adult immunization in general. That 38.1% of countries worldwide lack functional systems to deliver routine adult vaccinations has major implications for future SARS-CoV-2 vaccine deployment.

WHO immunization position papers are permissive toward national immunization programs for the adult vaccines we assessed, indicating that countries may choose to target adults for immunization, but no position paper mandates such programs. However, a 2017 WHO expert convening acknowledged that there were data gaps hindering adult vaccination program policy-making globally, particularly in LICs and LMICs [2]. Investment in studies to better measure disease burden and the potential impact of adult immunization programs are needed to inform the value proposition of adult immunization in low resource settings. Further, while global vaccine policy has often been driven by estimates of mortality prevention or cost-effectiveness, the full impact of adult vaccine-preventable diseases may be better understood by different metrics [2]. The impact on a community when older adults lose independence or functional capacity after a vaccine-preventable illness is difficult to assess, and the value of missed unpaid work in the home or as a child caregiver is seldom measured in economic analyses [17]. The presence of favourable global programs for adult immunization is not sufficient to advance vaccination programs if impact estimates undervalue older adults’ lives or their contributions.

While we do not necessarily advocate the expansion of routine adult immunization services into low- and low-middle income country settings, such programs can provide public health value. In addition to the direct impact of routine immunization, there are additional benefits of strong adult immunization programs. For example, they can provide platforms for the delivery of other preventive interventions and strengthen primary care [18]. They can enhance public health responses during public health emergencies requiring supplemental immunization activities, such as the current SARS-CoV-2 pandemic. In 2009, having a seasonal influenza vaccination program was significantly associated with the deployment of pandemic influenza vaccines when they were available [19]. This was attributed both to infrastructure preparedness as well as individual attitudes regarding vaccination, both of which are enhanced when a country has a functional adult influenza vaccination program [19]. Finally, strengthening countries’ capacities to provide immunization services across the life course should occur concurrent to the development of new vaccines targeting adult age groups, so that delivery systems will be in place once new vaccines become available.

A critical component to strong immunization systems is the NITAG, and we found an association between adult immunization programs in countries and functional NITAGs. Within an individual country, the NITAG plays a critical role in evaluating vaccines, interpreting available safety and efficacy data, and applying relevant data to policy recommendations most appropriate for a given country. NITAGs may consider targeting health care workers for influenza vaccination as the most logical first step to advancing immunization in LICs and LMICs. There are existing platforms to transport, store, and deliver vaccines within health care systems [20], the total population of health care workers is small compared to other influenza risk groups, and sensitizing health care workers to the benefits of vaccination has effects on their advocacy of vaccines to patients [21]. A healthy medical workforce is critical to ensuring that essential health care services continue during disease epidemics.

This study should be interpreted in the context of its strengths and limitations. To our knowledge, the JRF is the only global source of national immunization data and is therefore the only dataset that would permit a quantification of global adult immunization programs. The survey is a routine public health instrument that is completed annually by Ministries of Health with extensive quality checks at the regional and global levels. It can be an important platform to monitor adult vaccine policy development over time. However, the JRF relies on national self-reporting, and there may be errors introduced at the country level that would be difficult to identify or correct. These data capture only what is recommended at the government policy level, though may not describe vaccine use in the private sector. Further, the questions regarding the presence of adult immunization programs, particularly for PCV, require free-text responses for program target groups. WHO has moved to a web-based application to collect the 2019 JRF data, which will provide opportunities to improve the questionnaire and decrease potential reporting bias. We believe that engaging WHO Regional Office immunization focal points as a data quality check was a strength to this study, as they are highly informed about the routine immunization programs within their regions and are likely to know about any adult immunization programs, particularly in LICs and LMICs.

Finally, the presence of a national adult immunization program does not necessarily indicate substantial vaccine use. For example, while Estonia has an influenza vaccination program targeting older adults, it has reported only 2% vaccine coverage for this group in the 2016-2017 season [22]. In a 2015 review of global influenza vaccine distribution, the International Federation of Pharmaceutical Manufacturers and Associations estimated that about 95% of global influenza vaccine distribution occurs in the Americas, European, and Western Pacific Regions [23], despite 17 countries reporting adult influenza vaccination programs outside of those regions. While the JRF does collect information on influenza vaccination coverage of older adults and persons with chronic medical conditions, there is substantial data missingness making analyses of global influenza vaccine use in these risk groups infeasible [8]. Our analysis determined whether routine national programs are in place to immunize adults, but it could not ascertain whether systems are sufficient for SARS-CoV-2 vaccination campaigns.

If vaccines for SARS-CoV-2 become licensed, they will be deployed in many countries without adult immunization infrastructures. Substantial resources will be needed to ensure the equitable distribution of potentially life-saving vaccines to persons at high risk for severe disease and to protect essential workers. In 2020, the SARS-CoV-2 pandemic is disproportionately affecting older adults and is a threat to the safety of health care workers [24], yet nearly 40% of countries globally have no immunization infrastructures to provide adult vaccination, and nearly 60% of the world’s population lives in countries without routine adult immunization programs. Further, WHO advises the use of influenza vaccine, where feasible, for health care workers, older adults, and pregnant women during the SARS-CoV-2 pandemic [1]. The success of campaigns to deliver these vaccines is also threatened by lack of sufficient adult immunization infrastructures. Systems for social mobilization and outreach, vaccine storage and handling, delivery, and waste management for adult immunization do not exist in much of the world, and they will have to be developed to support the pandemic response. WHO is currently planning for an initial tranche of vaccines to cover 3% of country populations [25]. While this estimate will differ from country to country based on advanced market purchase agreements, the anticipated low supply of SARS-CoV-2 vaccines early after they gain licensure will limit stress on global immunization systems deploying them. Our study suggests that the global SARS-CoV-2 pandemic response should address disparities in public health resources now and strengthen immunization systems to maximize the impact and equity of future pandemic vaccine deployment.

## Data Availability

All data are available from the references provided.

http://www.who.int/immunization/monitoring_surveillance/routine/reporting/reporting/en/

https://datahelpdesk.worldbank.org/knowledgebase/articles/906519

https://databank.worldbank.org/reports.aspx?source=2&series=SH.XPD.GHED.PC.CD

## Notes

### Competing Interest Statement

The authors have declared no competing interest.

### Funding Statement

There was no specific funding for this study.

### Author Declarations

None needed, we analyzed publicly available dataset with country-level data.

### Summary of Updates

Added a European Region-specific analysis and minor changes of format, diction, and grammar for sense.

