## Supplement for "National Routine Adult Immunization Programs among World Health Organization Member States: An Assessment of Health Systems to Deploy Future SARS-CoV-2 Vaccines"

**DATA SUPPLEMENT**

**Supplemental Table 1. Countries Responding to Specific JRF Question Regarding Presence of an Adult Immunization Program, by Vaccine**

| WHO Region | Hepatitis B Vaccine | | Herpes Zoster Vaccine | | Influenza Vaccine | | Pneumococcal Conjugate Vaccine | | Pneumococcal Polysaccharide Vaccine | | Any of the Assessed Vaccines | |
|---|---|---|---|---|---|---|---|---|---|---|---|---|
| | n | % | n | % | n | % | n | % | n | % | n | % |
| Africa (n=47) | 3 | 6.4 | 0 | 0.0 | 3 | 6.4 | 41 | 87.2 | 0 | 0.0 | 41 | 87.2 |
| Americas (n=35) | 31 | 88.6 | 4 | 11.4 | 32 | 91.4 | 28 | 80.0 | 16 | 45.7 | 35 | 100.0 |
| Eastern Mediterranean (n=21) | 7 | 33.3 | 1 | 4.8 | 13 | 61.9 | 16 | 76.2 | 4 | 19.1 | 20 | 95.2 |
| Europe (n=53) | 37 | 69.8 | 31 | 58.5 | 50 | 94.3 | 48 | 90.6 | 34 | 64.2 | 52 | 98.1 |
| South-East Asia (n=11) | 0 | 0.0 | 0 | 0.0 | 1 | 9.1 | 5 | 45.5 | 0 | 0.0 | 6 | 54.6 |
| Western Pacific (n=27) | 5 | 18.5 | 2 | 7.4 | 15 | 55.6 | 19 | 70.4 | 5 | 18.5 | 22 | 81.5 |
| Worldwide (n=194) | 83 | 42.8 | 38 | 19.6 | 114 | 58.8 | 157 | 80.9 | 59 | 30.4 | 176 | 90.7 |

### Supplemental Table 2. Economic Characteristics of WHO Member States in 2018, by Reported National Adult Immunization Programs

| | HepB program | | | | | HZV program | | | | | Influenza vaccination program | | | | | PCV program | | | | | PPSV program | | | | |
|---|---|---|---|---|---|---|---|---|---|---|---|---|---|---|---|---|---|---|---|---|---|---|---|---|---|
| | Yes (N=72) | | No (N=121) | | p-value[1] | Yes (N=17) | | No (N=175) | | p-value | Yes (n=113) | | No (N=79) | | p-value | Yes (N=16) | | No (N=176) | | p-value | Yes (N=35) | | No (N=157) | | p-value |
| | n | % | n | % | | n | % | n | % | | n | % | n | % | | n | % | n | % | | n | % | n | % | |
| WB Income group[2] | | | | | | | | | | | | | | | | | | | | | | | | | |
| Low income (n=34) | 0 | 0.0 | 34 | 28.1 | <0.001 | 0 | 0.0 | 34 | 19.4 | <0.001 | 1 | 0.9 | 33 | 41.8 | <0.001 | 0 | 0.0 | 34 | 19.3 | <0.001 | 0 | 0.0 | 34 | 21.7 | <0.001 |
| Lower-middle income (n=46) | 7 | 9.9 | 39 | 32.1 | | 0 | 0.0 | 46 | 26.3 | | 15 | 13.3 | 31 | 39.2 | | 0 | 0.0 | 46 | 26.1 | | 2 | 5.7 | 44 | 28.0 | |
| Upper middle income (n=56) | 29 | 40.9 | 27 | 22.3 | | 1 | 5.9 | 55 | 31.4 | | 42 | 37.2 | 14 | 17.7 | | 0 | 0.0 | 56 | 31.8 | | 6 | 17.1 | 50 | 31.9 | |
| High income (n=56) | 35 | 49.3 | 21 | 17.4 | | 16 | 94.1 | 40 | 22.9 | | 55 | 48.7 | 1 | 1.3 | | 16 | 100.0 | 40 | 22.7 | | 27 | 77.1 | 29 | 18.5 | |
| Gavi eligible | Yes (N=72) | | No (N=123) | | p-value[1] | Yes (N=17) | | No (N=177) | | p-value | Yes (n=114) | | No (N=80) | | p-value | Yes (N=16) | | No (N=178) | | p-value | Yes (N=35) | | No (N=159) | | p-value |
| | n | % | n | % | | n | % | n | % | | n | % | n | % | | n | % | n | % | | n | % | n | % | |
| Yes (n=48) | 1 | 1.4 | 47 | 38.2 | <0.001 | 0 | 0.0 | 48 | 27.1 | 0.01 | 0 | 0.0 | 48 | 60.0 | <0.001 | 0 | 0.0 | 48 | 27.0 | 0.02 | 0 | 0.0 | 48 | 31.8 | <0.001 |
| No (n=146) | 70 | 98.6 | 76 | 61.8 | | 17 | 100.0 | 129 | 72.9 | | 114 | 100.0 | 32 | 40.0 | | 16 | 100.0 | 130 | 73.0 | | 43 | 100.0 | 103 | 68.2 | |

Abbreviations: HepB = Hepatitis B vaccine; HZV= Herpes Zoster Vaccine; PCV = pneumococcal conjugate vaccine; PPSV = pneumococcal polysaccharide vaccine; HPV= human papillomavirus vaccine

1. p-value for trend (World Bank income group comparisons)
2. Niue and The Cook Islands, not World Bank member countries, are excluded from the income categories
3. Reference category
4. Chi-square test (Gavi eligibility comparisonsf

**Supplemental Table 3. Median per Capita Health Care Expenditures of WHO Member States in 2018, by reported National Adult Immunization Program**

| **Adult Vaccination program** | **Has Program** | **Statistic** | **Median per capita health care expenditure (USD)[1]** |
|---|---|---|---|
| HepB program | Yes (N=68) | Median (IQR) | 884 (412, 2118) |
| | No (N=116) | Median (IQR) | 195 (45, 732) |
| | | **p-value[2]** | **<0.001** |
| HZV program | Yes (N=17) | Median (IQR) | 2,882 (1,845, 3,465) |
| | No (N=167) | Median (IQR) | 314 (62, 900) |
| | | **p-value** | **<0.001** |
| Influenza vaccination program | Yes (N=109) | Median (IQR) | 933 (482, 2,226) |
| | No (N=75) | Median (IQR) | 59 (25, 196) |
| | | **p-value** | **<0.001** |
| PCV program | Yes (N=16) | Median (IQR) | 2,221 (1335, 3510) |
| | No (N=168) | Median (IQR) | 319 (63, 884) |
| | | **p-value** | **<0.001** |
| PPSV program | Yes (N=43) | Median (IQR) | 1,676 (930, 3,331) |
| | No (N=151) | Median (IQR) | 208 (52, 564) |
| | | **p-value** | **<0.001** |

HepB= hepatitis B; HZV = Herpes zoster virus vaccine; PCV = pneumococcal conjugate vaccine; PPSV= pneumococcal polysaccharide vaccine

[1]Excluding 10 countries with missing health care expenditure data

[2]Kruskal-Wallis test for difference in medians

### Supplemental Table 4. Immunization Program Characteristics of WHO Member states in 2018, by reported National Adult Immunization Programs

| | | HepB program | | | | | HZV Program | | | | | Influenza vaccine program | | | | | PCV program | | | | | PPSV program | | | | |
|---|---|---|---|---|---|---|---|---|---|---|---|---|---|---|---|---|---|---|---|---|---|---|---|---|---|---|
| Introduced Hepatitis B birth dose | n | Yes | | No | | p-value | Yes | | No | | p-value | Yes | | No | | p-value | Yes | | No | | p-value | Yes | | No | | p-value |
| | | n | % | n | % | | n | % | n | % | | n | % | n | % | | n | % | n | % | | n | % | n | % | |
| Yes | 128 | 53 | 41.4 | 75 | 58.6 | 0.05 | 14 | 10.9 | 114 | 89.1 | 0.14 | 91 | 71.1 | 37 | 28.9 | <0.001 | 12 | 9.4 | 116 | 90.6 | 0.43 | 27 | 21.1 | 101 | 78.9 | 0.12 |
| No | 66 | 18 | 27.3 | 48 | 72.7 | | 3 | 4.6 | 63 | 95.5 | | 23 | 34.9 | 43 | 65.2 | | 4 | 6.1 | 62 | 93.9 | | 8 | 12.1 | 58 | 87.9 | |
| Introduced HPV | | | | | | | | | | | | | | | | | | | | | | | | | | |
| Yes | 90 | 50 | 55.6 | 40 | 44.4 | <0.001 | 14 | 10.9 | 114 | 89.1 | <0.001 | 73 | 81.1 | 17 | 18.9 | <0.001 | 13 | 14.4 | 77 | 85.6 | <0.01 | 29 | 32.2 | 61 | 67.8 | <0.001 |
| No | 104 | 21 | 20.2 | 83 | 79.8 | | 17 | 18.9 | 73 | 81.1 | | 41 | 39.4 | 63 | 60.6 | | 3 | 2.9 | 101 | 97.1 | | 6 | 5.8 | 98 | 94.2 | |
| Introduced Rotavirus vaccine | | | | | | | | | | | | | | | | | | | | | | | | | | |
| Yes | 101 | 40 | 39.6 | 61 | 60.4 | 0.37 | 13 | 12.9 | 88 | 87.1 | 0.04 | 56 | 55.5 | 45 | 44.6 | 0.33 | 10 | 9.9 | 91 | 90.1 | 0.38 | 23 | 22.8 | 78 | 77.2 | 0.07 |
| No | 93 | 31 | 33.3 | 62 | 66.7 | | 4 | 4.3 | 89 | 95.7 | | 58 | 62.4 | 35 | 37.6 | | 6 | 6.5 | 87 | 93.6 | | 12 | 12.9 | 81 | 87.1 | |
| Functioning NITAG[2] | | | | | | | | | | | | | | | | | | | | | | | | | | |
| Yes | 114 | 47 | 41.2 | 67 | 58.8 | 0.11 | 14 | 12.3 | 100 | 87.7 | 0.04 | 78 | 68.4 | 36 | 31.6 | <0.01 | 13 | 11.4 | 101 | 88.6 | 0.06 | 23 | 20.2 | 91 | 79.8 | 0.36 |
| No | 80 | 24 | 30.0 | 56 | 70.0 | | 3 | 3.8 | 77 | 96.3 | | 36 | 45.0 | 44 | 55.0 | | 3 | 3.8 | 77 | 96.3 | | 12 | 15.0 | 68 | 85.0 | |
| Eliminated maternal and neonatal tetanus | | | | | | | | | | | | | | | | | | | | | | | | | | |
| Yes | 180 | 71 | 39.4 | 109 | 60.0 | <0.01 | 17 | 9.4 | 163 | 90.6 | 0.23 | 114 | 63.3 | 66 | 36.7 | <0.001 | 16 | 8.9 | 164 | 91.1 | 0.24 | 35 | 19.4 | 145 | 80.6 | 0.07 |
| No | 14 | 0 | 0.0 | 14 | 100.0 | | 0 | 0.0 | 14 | 100.0 | | 0 | 0.0 | 14 | 100.0 | | 0 | 0.0 | 14 | 100.0 | | 0 | 0.0 | 14 | 100.0 | |
| DTP coverage ≥95% nationally | | | | | | | | | | | | | | | | | | | | | | | | | | |
| Yes | 84 | 39 | 46.4 | 45 | 53.6 | 0.01 | 7 | 8.3 | 77 | 91.7 | 0.85 | 62 | 73.8 | 22 | 26.2 | <0.001 | 10 | 11.9 | 74 | 88.1 | <0.01 | 18 | 21.4 | 66 | 78.6 | 0.28 |
| No | 110 | 32 | 29.1 | 78 | 70.9 | | 10 | 9.1 | 100 | 90.9 | | 52 | 47.3 | 58 | 52.7 | | 6 | 5.5 | 104 | 94.6 | | 17 | 15.5 | 93 | 84.6 | |

HepB = Hepatitis B vaccine; HZV= Herpes Zoster Vaccine; PCV = pneumococcal conjugate vaccine; PPSV = pneumococcal polysaccharide vaccine; HPV= human papillomavirus vaccine; DTP= diphtheria, tetanus and pertussis containing vaccine

1. Chi-square test
2. National Immunization Technical Advisory Group; limited to n=134 countries for which this information was available

**Supplemental Table 5A. Analysis of Immunization Program Characteristics associated with the presence of one or more adult vaccination programs**

| | Any adult vaccination program | | |
|---|---|---|---|
| | Yes (N=120) | | |
| | n | % | aOR[1] (95% CI) |
| Introduced Hepatitis B birth dose | | | |
| Yes | 94 | 78.3 | 3.26 (1.2, 9.0) |
| No | 26 | 21.7 | |
| Introduced HPV | | | |
| Yes | 76 | 63.3 | 3.48 (1.3, 9.4) |
| No | 44 | 36.7 | |
| Introduced Rotavirus vaccine | | | |
| Yes | 60 | 50.0 | 1.13 (0.4, 2.9) |
| No | 60 | 50.0 | |
| Functional NITAG[2] | | | |
| Yes | 82 | 68.3 | 6.74 (2.3, 19.7) |
| No | 38 | 31.7 | |
| Eliminated maternal and neonatal tetanus | | | |
| Yes | 120 | 100.0 | n/a |
| No | 0 | 0.0 | |
| DTP coverage ≥95% nationally | | | |
| Yes | 66 | 55.0 | 1.57 (0.6, 3.9) |
| No | 54 | 45.0 | |
| HIC or UMIC2 | | | |
| Yes | 100 | 84.0 | 19.28 (6.5, 57.7) |
| No | 19 | 16.0 | |

1. Adjusted for all other characteristics in the table
2. Two countries without world bank income classifications excluded from denominator

**Supplemental Table 5B. Analysis of Immunization Program Characteristics associated with the presence of individual adult vaccination programs**

| | Adult HepB program | | | Adult HZV Program | | | Adult influenza vaccination program | | | Adult PCV Program | | | Adult PPSV Program | | |
|---|---|---|---|---|---|---|---|---|---|---|---|---|---|---|---|
| | Yes (N=71) | | | Yes (N=17) | | | Yes (N=114) | | | Yes (N=16) | | | Yes (N=35) | | |
| | n | % | aOR (95% CI) | n | % | aOR (95% CI) | n | % | aOR[1] (95% CI) | n | % | aOR (95% CI) | n | % | aOR (95% CI) |
| Introduced Hepatitis B birth dose | | | | | | | | | | | | | | | |
| Yes | 53 | 74.7 | 1.0 (0.4, 2.3) | 14 | 82.4 | 2.0 (0.5, 8.5) | 91 | 79.8 | 3.7 (1.4, 10.1) | 12 | 75.0 | 0.9 (0.2, 3.1) | 27 | 77.1 | 1.3 (0.5, 3.4) |
| No | 18 | 25.4 | | 3 | 17.7 | | 23 | 20.2 | | 4 | 25.0 | | 8 | 22.9 | |
| Introduced HPV | | | | | | | | | | | | | | | |
| Yes | 50 | 70.4 | 1.9 (0.9, 4.1) | 17 | 100.0 | n/a | 73 | 64.0 | 3.4 (1.3, 9.0) | 13 | 81.3 | 1.9 (0.4, 7.8) | 29 | 82.9 | 3.3 (1.2, 9.3) |
| No | 21 | 29.6 | | 0 | 0.0 | | 41 | 36.0 | | 3 | 18.8 | | 6 | 17.1 | |
| Introduced Rotavirus vaccine | | | | | | | | | | | | | | | |
| Yes | 40 | 56.3 | 1.8 (0.9, 3.9) | 13 | 76.5 | 2.5 (0.7, 9.6) | 56 | 49.1 | 0.95 (0.4, 2.4) | 10 | 62.5 | 2.1 (0.6, 7.1) | 23 | 65.7 | 2.18 (0.9, 5.3) |
| No | 31 | 43.7 | | 4 | 23.5 | | 58 | 50.9 | | 6 | 37.5 | | 12 | 34.3 | |
| Functional NITAG[2] | | | | | | | | | | | | | | | |
| Yes | 47 | 66.2 | 1.5 (0.7, 3.1) | 14 | 82.4 | 3.17 (0.8, 12.8) | 78 | 68.4 | 5.6 (2.1, 15.3) | 13 | 81.3 | 3.1 (0.8, 11.9) | 23 | 65.7 | 1.2 (0.5, 2.8) |
| No | 24 | 33.8 | | 3 | 17.7 | | 36 | 31.6 | | 3 | 18.8 | | 12 | 34.3 | |
| Eliminated maternal and neonatal tetanus | | | | | | | | | | | | | | | |
| Yes | 71 | 100.0 | n/a | 17 | 100.0 | n/a | 114 | 100.0 | n/a | 16 | 100.0 | n/a | 35 | 100.0 | n/a |
| No | 0 | 0.0 | | 0 | 0.0 | | 0 | 0.0 | | 0 | 0.0 | | 0 | 0.0 | |
| DTP coverage ≥95% nationally | | | | | | | | | | | | | | | |

| | | | | | | | | | | | | | | | |
|---|---|---|---|---|---|---|---|---|---|---|---|---|---|---|---|
| Yes | 39 | 54.9 | 1.2 (0.6, 2.1) | 7 | 41.2 | 0.7 (0.2, 2.3) | 62 | 54.4 | 1.2 (0.5, 2.8) | 10 | 62.5 | 1.7 (0.5, 5.5) | 18 | 51.4 | 1.0 (0.4, 2.3) |
| No | 32 | 45.1 | | 10 | 58.8 | | 52 | 45.6 | | 6 | 37.5 | | 17 | 48.6 | |
| HIC or UMIC[2] | | | | | | | | | | | | | | | |
| Yes | 64 | 90.1 | 10.1 (3.6,28.1) | 17 | 100.0 | n/a | 97 | 85.8 | 17.3 (6.2, 48.4) | 16 | 100.0 | n/a | 33 | 94.3 | 9.2 (1.9, 45.1) |
| No | 7 | 9.9 | | 0 | 0.0 | | 16 | 14.2 | | 0 | 0.0 | | 2 | 5.7 | |

1. Adjusted for all other characteristics in the table
2. Two countries without world bank income classifications excluded from denominator
